# Diagnostic Accuracy of Dynamic Supine-to-Sitting Radiography for Acute Osteoporotic Vertebral Fractures. A Preliminary Single-Center Diagnostic Accuracy Study

**DOI:** 10.64898/2026.08.06.26359902

**Authors:** Ryota Kimura, Norio Yamamoto, Kazuma Doi

## Abstract

**Background:** Acute osteoporotic vertebral fractures (OVFs) may be difficult to detect on conventional radiographs, particularly before substantial vertebral collapse occurs. Comparing supine and sitting lateral radiographs may reveal load-dependent vertebral mobility. This preliminary study evaluated the diagnostic accuracy of supine-to-sitting dynamic radiography for detecting MRI-confirmed acute OVFs.

**Methods:** This retrospective, single-center diagnostic accuracy study included consecutive patients who underwent paired supine and sitting lateral radiography and MRI of the same spinal region between April 2024 and July 2026. Dynamic radiographs were interpreted by a board-certified orthopedic and spine surgeon who was blinded to the MRI findings. MRI was independently interpreted by a second board-certified orthopedic surgeon and served as the reference standard. The primary outcome was patient-level sensitivity and specificity. Vertebra-level diagnostic accuracy was evaluated secondarily, with patient-cluster bootstrap confidence intervals used to account for within-patient correlation.

**Results:** Sixty-three patients (mean age, 80.6 ± 9.0 years; 51 women [81.0%]) and 490 evaluable vertebrae were analyzed. MRI identified acute OVFs in 34 patients and 36 vertebrae. At the patient level, dynamic radiography yielded 31 true-positive, no false-positive, three false-negative, and 29 true-negative results. Sensitivity was 91.2% (95% confidence interval [CI], 76.3%–98.1%), specificity was 100.0% (95% CI, 88.1%–100.0%), positive predictive value was 100.0%, negative predictive value was 90.6%, and overall accuracy was 95.2%. At the vertebral level, sensitivity was 91.7% (33/36; patient-cluster bootstrap 95% CI, 81.3%–100.0%) and specificity was 100.0% (454/454). The three missed fractures involved T9, L2, and L3. No false-positive vertebrae were observed.

**Conclusions:** Supine-to-sitting dynamic radiography demonstrated high patient-level sensitivity and no observed false-positive findings for MRI-confirmed acute OVFs. It may provide a practical complementary diagnostic option when MRI is not immediately available. However, a negative dynamic radiographic examination does not exclude an acute fracture, and the apparent perfect specificity requires validation in larger, prospective multi-reader studies.

## Introduction

Osteoporotic vertebral fractures (OVFs) are common fragility fractures in older adults and are associated with pain, impaired physical function, reduced health-related quality of life, subsequent fractures, and increased mortality.^1,2^ Timely recognition of an acute OVF is clinically important because the diagnosis affects pain management, mobilization, bracing, osteoporosis treatment, and decisions regarding further intervention. Nevertheless, acute OVFs can be difficult to identify on conventional radiographs, particularly when vertebral collapse is minimal or when chronic deformities coexist.

Magnetic resonance imaging (MRI) can demonstrate fracture-related bone marrow edema and is widely used as the reference standard for determining fracture acuity.^3^ However, MRI may not be immediately available, may require a separate appointment, and cannot be performed in some patients. Plain radiography is inexpensive and widely accessible, but a supine image may underestimate an unstable fracture because unloading can partially restore vertebral body height. Comparing lateral radiographs obtained under non-weight-bearing and loaded conditions may therefore reveal fracture mobility that is not apparent on a single image.

Previous studies have evaluated positional changes in vertebral body height using supine and sitting or other loaded radiographs. Niimi et al. reported that mobility assessed by paired supine and sitting lateral radiographs improved identification of acute OVFs compared with either position alone.^4^ More recent pilot and prospective work has likewise suggested that positional radiography may provide useful diagnostic information when MRI is not readily available.^5,6^ However, the evidence remains sparse and heterogeneous. Several studies enrolled only patients with reference-standard-positive fractures or did not provide complete 2 x 2 data, limiting estimation of both sensitivity and specificity. In addition, vertebra-level analyses require methods that account for the correlation of multiple vertebrae within the same patient.

The primary aim of this preliminary study was to estimate the patient-level diagnostic accuracy of dynamic lateral radiography, defined as a comparison between supine and sitting positions, for detecting acute OVFs using MRI as the reference standard. Secondary aims were to estimate vertebra-level diagnostic accuracy and to describe correct localization of MRI-positive vertebral levels. This preliminary phase uses a single expert radiographic reader and is intended to inform a subsequent, adequately sized multi-reader study evaluating interobserver agreement across readers with different levels of orthopedic training.

## Methods

### Study design and reporting

This retrospective, single-center diagnostic accuracy study included consecutive patients evaluated between April 2024 and July 2026. The index test was paired lateral radiography obtained in the supine and sitting positions, and the reference standard was MRI. Reporting followed the Standards for Reporting Diagnostic Accuracy Studies (STARD) 2015 statement.^7^ This analysis constitutes a preliminary phase of a planned larger multi-reader investigation; interobserver agreement was not evaluated in the present phase.

### Participants

Patients were eligible if they (1) were aged 60 years or older; (2) presented with acute thoracolumbar or low back pain of no more than 3 weeks’ duration after low-energy trauma or without an identifiable traumatic event; (3) were clinically suspected of having an acute OVF; (4) underwent both supine and sitting lateral radiography; and (5) underwent MRI covering the same spinal region within 14 days of radiography. At least one vertebral body had to be evaluable on both radiographic views and MRI. Patients were excluded for high-energy trauma, suspected or confirmed spinal infection, malignant or other pathological vertebral fracture, a new traumatic event between the index test and MRI, or imaging of insufficient quality to determine the index-test or reference-standard result. Prior vertebral fractures and spinal instrumentation were not automatic exclusions unless they prevented evaluation of the relevant vertebral level. Patients unable to tolerate the sitting position were recorded separately in the participant flow and were not silently removed from the source cohort.

### Index test: dynamic supine-to-sitting radiography

Lateral radiographs were obtained in the supine and unsupported or supported sitting positions during the same clinical episode. Radiographs were obtained as part of routine clinical practice according to the institutional imaging protocol. Minor variations in sitting posture and the use of support were permitted according to pain severity and physical function. Only vertebral levels visible with diagnostic quality on both views were considered evaluable.

The paired radiographs were assessed level by level. An index-test-positive vertebra was defined by a discernible reduction in vertebral body height or an increase in wedge or compression deformity on the sitting image compared with the supine image, consistent with positional mobility of the vertebral body.^4^ No post hoc threshold was selected from the MRI results. If a quantitative height-change threshold is used in the final dataset, its definition and measurement landmarks will be specified before analysis.

The preliminary readings were performed by one board-certified orthopedic surgeon and certified spine surgeon with 14 years of clinical experience in orthopedic surgery and 10 years of experience in spine surgery, including routine interpretation of radiographs in patients with osteoporotic vertebral fractures. No formal calibration session was performed because only one experienced reader participated in this preliminary study. For the study assessment, radiographs were de-identified and reviewed in a prespecified order without access to MRI findings. Radiographic judgments were recorded and locked before the reference-standard assessment was accessed. The reader was permitted to know the patient’s age and the imaged spinal region but not the MRI result or final clinical diagnosis. Any deviation from this blinding procedure will be reported explicitly.

### Reference standard

MRI served as the reference standard for fracture acuity. The MRI protocol included sagittal T1-weighted, T2-weighted, and short-tau inversion recovery (STIR) sequences. An acute OVF was defined as a vertebral body demonstrating a marrow edema pattern compatible with acute fracture, typically low signal intensity on T1-weighted images and high signal intensity on STIR images, with or without an associated fracture line or vertebral deformity.^3^ A deformed vertebra without marrow edema was classified as a prior or chronic vertebral fracture.

MRI examinations were independently interpreted by a second board-certified orthopedic surgeon who did not participate in the interpretation of the dynamic radiographs and was blinded to the index-test results. The reference-standard result was recorded separately for every evaluable vertebral level.

### Data collection and analysis units

The following patient-level variables were collected: age, sex, height, weight, body mass index, prior vertebral fracture, and the number of vertebral bodies evaluable on both radiography and MRI. Index-test and MRI results were recorded for each vertebral level. For the primary patient-level analysis, a patient was considered evaluable when at least one vertebral level was visible with diagnostic quality on both radiographic views and MRI. For the secondary vertebra-level analysis, a vertebra was included only when the same level was evaluable on the supine radiograph, sitting radiograph, and MRI. The reason for every non-evaluable level or examination was documented.

The primary unit of analysis was the patient. The index test was classified as positive when at least one vertebra was positive on dynamic radiography, and the reference standard was classified as positive when at least one acute OVF was present on MRI. The secondary unit of analysis was the vertebral body. Each evaluable vertebra was cross-classified as true positive, false positive, false negative, or true negative by comparing the level-specific dynamic-radiograph result with the level-specific MRI result. Among MRI-positive patients, correct localization was summarized separately as the proportion in whom at least one radiograph-positive vertebral level matched an MRI-positive level.

### Outcomes

The primary outcome was patient-level diagnostic accuracy, summarized by sensitivity and specificity. Secondary diagnostic performance measures included patient-level positive and negative predictive values, overall accuracy, positive and negative likelihood ratios, vertebra-level sensitivity and specificity, other vertebra-level diagnostic performance measures, and the correct-level localization proportion. The frequency of non-evaluable sitting examinations and non-evaluable vertebral levels was also reported.

### Sample size

No formal hypothesis-testing sample-size calculation was performed for this preliminary study. All eligible patients available during the prespecified study period were included. Estimates were interpreted primarily through their 95% confidence intervals rather than statistical significance. The observed prevalence, precision, non-evaluable rate, and within-patient clustering will be used to refine the sample-size calculation for the subsequent multi-reader study.

### Statistical analysis

Continuous variables were summarized as mean and standard deviation or median and interquartile range according to their distribution; categorical variables were summarized as counts and percentages. Sensitivity, specificity, predictive values, accuracy, and likelihood ratios were calculated from 2 x 2 contingency tables with 95% confidence intervals. Because the index test was binary and no threshold was varied, receiver operating characteristic curves and area under the curve were not calculated.

For the primary patient-level analysis, exact Clopper-Pearson 95% confidence intervals were calculated for sensitivity, specificity, predictive values, and accuracy. For the secondary vertebra-level analysis, correlation among multiple vertebrae from the same patient was addressed using a patient-cluster bootstrap: patients, rather than individual vertebrae, were resampled with replacement, and all evaluable vertebrae belonging to a selected patient were retained in each replicate.^8^ Percentile 95% confidence intervals were obtained from 10,000 bootstrap replicates. Exact binomial intervals treating vertebrae as independent were also reported for comparability with previous studies. If a zero cell prevented calculation of a likelihood ratio, the measure was reported as not estimable; a continuity-corrected estimate was presented only as a sensitivity analysis.

The primary patient-level analysis included all evaluable patients without imputation. The secondary vertebra-level analysis used available evaluable levels without imputation. Missing, indeterminate, and technically non-evaluable results were described explicitly. Sensitivity analyses were planned (1) excluding patients with prior vertebral fractures and, if the levels of old fractures were available, excluding old-fracture levels; and (2) treating indeterminate index-test results as incorrect classifications. All analyses were performed using Python version 3.12.13 (Python Software Foundation, Wilmington, DE, USA), with NumPy version 2.3.5 and SciPy version 1.17.0. All statistical tests were two-sided with a significance level of 0.05; no multiplicity-adjusted confirmatory testing was planned because the study was exploratory.

### Ethics

The study was conducted in accordance with the Declaration of Helsinki and was approved by the Ethics Committee of Kitaakita Municipal Hospital (approval no. 72). Given the retrospective nature of the study, written informed consent was not obtained individually. Instead, information about the study was publicly disclosed, and patients were provided with the opportunity to opt out of participation.

## Results

### Study population

A total of 63 patients who underwent both dynamic radiography in the supine and sitting positions and magnetic resonance imaging (MRI) were included in the analysis. The mean age was 80.6 ± 9.0 years (range, 60-98 years), and 51 patients (81.0%) were women. The mean body mass index was 22.5 ± 4.2 kg/m2. Twelve patients (19.0%) had at least one prior vertebral fracture. The median interval between dynamic radiography and MRI was 0 days (interquartile range [IQR], 0-3 days; range, 0-14 days), and both examinations were performed on the same day in 33 patients (52.4%). Baseline characteristics are summarized in Table 1.

**Table 1.** Participant and imaging characteristics.

| Characteristic | Overall (N = 63) |
| --- | --- |
| Age, years | 80.6 ± 9.0 |
| Female sex, n (%) | 51 (81.0%) |
| Height, cm | 152.4 ± 8.2 |
| Weight, kg | 52.6 ± 11.8 |
| BMI, kg/m<sup>2</sup> | 22.5 ± 4.2 |
| Prior vertebral fracture present, n (%) | 12 (19.0%) |
| Number of prior vertebral fractures, median (IQR) | 0 (0-0) |
| X-ray-MRI interval, days, median (IQR) | 0 (0-3) |
| Same-day X-ray and MRI, n (%) | 33 (52.4%) |
| Assessable vertebrae per patient | 7.8 ± 0.8 |
| MRI-positive patients, n (%) | 34 (54.0%) |
| MRI-positive vertebrae, n (%) | 36 (7.3%) |

Values are mean +/- standard deviation, median (interquartile range), or n (%), as indicated. BMI, body mass index; MRI, magnetic resonance imaging.

A total of 490 vertebrae were jointly assessable on dynamic radiography and MRI, corresponding to a mean of 7.8 ± 0.8 vertebrae per patient (range, 6-10 vertebrae). MRI identified acute vertebral fractures in 34 patients (54.0%) and 36 vertebrae (7.3%). Dynamic radiography was positive in 31 patients (49.2%) and 33 vertebrae (6.7%).

### Patient-level diagnostic accuracy

In the primary per-patient analysis, there were 31 true-positive, 0 false-positive, 3 false-negative, and 29 true-negative patients (Table 2). Sensitivity was 91.2% (95% confidence interval [CI], 76.3%-98.1%), specificity was 100.0% (95% CI, 88.1%-100.0%), positive predictive value was 100.0% (95% CI, 88.8%-100.0%), negative predictive value was 90.6% (95% CI, 75.0%-98.0%), and overall accuracy was 95.2% (95% CI, 86.7%-99.0%) (Table 3).

**Table 2.** Patient-level 2 x 2 contingency table using MRI as the reference standard.

| **Dynamic radiography** | **MRI positive** | **MRI negative** |
| --- | --- | --- |
| Positive | 31 | 0 |
| Negative | 3 | 29 |

MRI, magnetic resonance imaging.

**Table 3.** Patient-level diagnostic accuracy of dynamic radiography.

| **Metric** | **Estimate, %** | **95% CI, %** | **Numerator/denominator** |
| --- | --- | --- | --- |
| Sensitivity | 91.2 | 76.3-98.1 | 31/34 |
| Specificity | 100.0 | 88.1-100.0 | 29/29 |
| Positive predictive value | 100.0 | 88.8-100.0 | 31/31 |
| Negative predictive value | 90.6 | 75.0-98.0 | 29/32 |
| Accuracy | 95.2 | 86.7-99.0 | 60/63 |

CI, confidence interval. Exact 95% CIs were calculated using the Clopper-Pearson method.
Cohen's kappa was 0.905 (bootstrap 95% CI, 0.780-1.000), and the exact McNemar p value was 0.250.

Agreement between dynamic radiography and MRI at the patient level was high, with a Cohen’s kappa coefficient of 0.905 (bootstrap 95% CI, 0.780-1.000). The exact McNemar test did not demonstrate a statistically significant difference in the proportion of positive results between dynamic radiography and MRI (p = 0.250).

### Vertebral-level diagnostic accuracy

In the secondary per-vertebra analysis, the 2 x 2 contingency table comprised 33 true-positive, 0 false-positive, 3 false-negative, and 454 true-negative vertebrae (Table 4). Dynamic radiography demonstrated a sensitivity of 91.7% (95% CI, 77.5%-98.2%) and a specificity of 100.0% (95% CI, 99.2%-100.0%). The positive predictive value was 100.0% (95% CI, 89.4%-100.0%), the negative predictive value was 99.3% (95% CI, 98.1%-99.9%), and overall accuracy was 99.4% (95% CI, 98.2%-99.9%) (Table 5). Agreement between dynamic radiography and MRI was high, with a Cohen’s kappa coefficient of 0.953.

**Table 4.** Vertebral-level 2 x 2 contingency table using MRI as the reference standard.

| **Dynamic radiography** | **MRI positive** | **MRI negative** |
| --- | --- | --- |
| Positive | 33 | 0 |
| Negative | 3 | 454 |

MRI, magnetic resonance imaging.

**Table 5.** Vertebral-level diagnostic accuracy of dynamic radiography.

| **Metric** | **Estimate, %** | **95% CI, %** | **Numerator/denominator** |
| --- | --- | --- | --- |
| Sensitivity | 91.7 | 77.5-98.2 | 33/36 |
| Specificity | 100.0 | 99.2-100.0 | 454/454 |
| Positive predictive value | 100.0 | 89.4-100.0 | 33/33 |
| Negative predictive value | 99.3 | 98.1-99.9 | 454/457 |
| Accuracy | 99.4 | 98.2-99.9 | 487/490 |

CI, confidence interval. Exact 95% CIs were calculated using the Clopper-Pearson method.
These exact intervals treat vertebrae as independent; patient-cluster bootstrap intervals are reported in the text.

In the patient-cluster bootstrap analysis, the 95% CI was 81.3%-100.0% for sensitivity, 98.5%-100.0% for the negative predictive value, 98.6%-100.0% for accuracy, and 0.890-1.000 for Cohen’s kappa. Specificity was 100.0% in every bootstrap resample because no false-positive vertebrae were observed; therefore, the bootstrap distribution for specificity was degenerate.

### Discordant findings

All three discordant cases were MRI-positive and dynamic-radiography-negative. The missed fracture levels were T9, L2, and L3, with one missed fracture at each level. All three patients underwent both examinations on the same day. No false-positive vertebrae were observed, and no dynamic-radiography-positive case showed disagreement with MRI regarding the identified fracture level.

## Discussion

This preliminary diagnostic accuracy study found that dynamic radiography comparing supine and sitting positions had high diagnostic performance for MRI-confirmed acute osteoporotic vertebral fractures. In the primary patient-level analysis, sensitivity was 91.2% and specificity was 100.0%. In the secondary vertebra-level analysis, sensitivity was 91.7% and specificity was 100.0%. Agreement with MRI was correspondingly high at both analysis levels. The principal discordance was false negativity: dynamic radiography missed three MRI-positive patients and three MRI-positive vertebrae, whereas no false-positive finding was observed.

The high specificity is clinically plausible because the index test required a positional change in vertebral height or morphology between the unloaded supine position and the loaded sitting position. Such mobility is a relatively direct manifestation of mechanical instability at an acute fracture site. Conversely, the three false-negative cases indicate that not all MRI-positive fractures demonstrate sufficient mobility or morphologic change to be recognized on dynamic radiographs. Early fractures without measurable collapse, fractures with limited mobility, and vertebral levels that are difficult to visualize may remain radiographically occult. Because all three false-negative patients underwent MRI and radiography on the same day, the discordance is unlikely to be explained solely by interval progression between examinations.

Previous studies have generally supported the diagnostic value of positional or loading-dependent assessment, although reported accuracy has varied. Niimi et al. showed that evaluation of vertebral mobility using dynamic radiographs was more useful than assessment of supine or sitting radiographs alone.^4^ Based on the extractable vertebral-level 2 x 2 data from that study, sitting radiographs had a sensitivity of approximately 39.6% and a specificity of 94.4%. A recent prospective preprint by Yamamoto et al. reported a sensitivity of 77% and a specificity of 86% for sitting radiographs at the patient level.^6^

The higher sensitivity and specificity observed in the present cohort should not be interpreted as evidence that this protocol is inherently superior to those used in previous studies. Diagnostic accuracy is affected by the spectrum of disease, recruitment setting, fracture severity, definition of a positive dynamic finding, evaluable spinal levels, image-acquisition technique, and reader expertise. The MRI-positive prevalence in the present cohort was 54.0% at the patient level, indicating a clinically enriched population with a high pretest probability of fracture. A strict criterion requiring concordance at the same vertebral level may also have contributed to the absence of false-positive findings. Direct comparison among studies therefore requires harmonized eligibility criteria, imaging protocols, reader blinding, and analysis units.

These findings suggest that supine-to-sitting dynamic radiography may be a useful complementary test when MRI cannot be obtained immediately. In this cohort, a positive dynamic radiographic finding was highly concordant with MRI and may therefore provide timely evidence supporting initiation of fracture management. The technique is inexpensive, widely available, and can be incorporated into routine radiographic workflows without specialized equipment.

However, dynamic radiography should not be considered a replacement for MRI. Three of 34 MRI-positive patients had negative dynamic radiographs, corresponding to a patient-level false-negative rate of 8.8%. Thus, a negative examination does not fully exclude an acute fracture when clinical suspicion remains high. MRI remains particularly important when pain is severe or persistent, when the suspected level is not clearly visualized, when multiple or atypical lesions are possible, or when infection or malignancy must be excluded. The observed 100% specificity and positive predictive value are encouraging, but the lower confidence limits and absence of false-positive events require cautious interpretation.

This study has several strengths. First, both MRI-positive and MRI-negative patients were included, allowing construction of complete 2 x 2 contingency tables and estimation of both sensitivity and specificity. Second, the patient-level analysis was specified as the primary analysis because it directly reflects the clinical question of whether a patient has at least one acute fracture; the secondary vertebra-level analysis additionally assessed localization of the responsible fracture. Third, the imaging interval was short, with a median of 0 days and same-day imaging in more than half of the cohort, limiting misclassification caused by temporal changes in fracture morphology. Fourth, dynamic radiographs were interpreted without knowledge of the MRI findings, reducing the risk of review bias.

Several limitations should also be acknowledged. First, this was a retrospective, single-center, preliminary study with a modest sample size. The analysis included only patients who underwent both dynamic radiography and MRI, which may have introduced selection bias and may limit applicability to patients who cannot tolerate the sitting position or undergo MRI. Second, radiographic interpretation was performed by a single spine specialist. Although the dynamic radiographs were interpreted while blinded to the MRI findings, the estimates may reflect expert-reader performance, and interobserver reliability could not be assessed. Third, the primary analysis included only 34 MRI-positive and 29 MRI-negative patients, and no false-positive event occurred. Consequently, the patient-level point estimates of specificity and positive predictive value reached 100%, but these values should not be interpreted as proof of perfect performance. The secondary analysis included only 36 MRI-positive vertebrae. Fourth, multiple vertebrae were nested within individual patients. The exact binomial confidence intervals treat vertebrae as independent and may therefore be overly narrow. We addressed this issue with a patient-cluster bootstrap analysis, although the specificity interval remained degenerate because no false-positive event was observed. Fifth, femoral bone mineral density data were unavailable, limiting quantitative characterization of the underlying osteoporosis. Finally, the maximum interval between examinations was 14 days, and the cohort was drawn from a single Japanese institution; changes in fracture morphology over time and differences in patient spectrum or radiographic technique may limit generalizability.

## Conclusion

In this preliminary cohort, supine-to-sitting dynamic radiography demonstrated high patient-level sensitivity and no observed false-positive findings for MRI-confirmed acute osteoporotic vertebral fractures. Secondary vertebra-level analysis yielded similarly high diagnostic performance. The method may provide a practical complementary diagnostic option when MRI is not immediately available. Nevertheless, negative dynamic radiographs did not exclude acute fracture, and the 100% specificity estimate requires external validation. Larger prospective studies using blinded independent readers and formal interobserver comparisons are warranted.

## Data Availability Statement

The individual-level data generated and analyzed during this study are not publicly available because they contain potentially identifiable clinical information and their public disclosure was not covered by the ethics approval. De-identified aggregate data may be available from the corresponding author upon reasonable request.

